# A distinct metabolic profile associated with a fatal outcome in COVID-19 patients during early epidemic in Italy

**DOI:** 10.1101/2021.04.13.21255117

**Authors:** Elisa Saccon, Alessandra Bandera, Mariarita Sciumè, Flora Mikeloff, Abid Ali Lashari, Stefano Aliberti, Michael Sachs, Filippo Billi, Francesco Blasi, Erin Gabriel, Giorgio Costantino, Pasquale De Roberto, Shuba Krishnan, Andrea Gori, Flora Peyvandi, Luigia Scudeller, Christian L. Lorson, Luca Valenti, Kamal Singh, Luca Baldini, Nicola Stefano Fracchiolla, on behalf of the “COVID-19 NETWORK” working group, Ujjwal Neogi

## Abstract

Leveraging the unique biological resource based upon the initial COVID-19 patients in Policlinico di Milano (Italy), our study provides the first metabolic profile associated with a fatal outcome. The identification of potential predictive biomarkers offers a vital opportunity to employ metabolomics in a clinical setting as diagnostic tool of disease prognosis upon hospital admission.

## Introduction

As of February 2021, more than 100 million cases of coronavirus disease 2019 (COVID-19), caused by the novel severe acute respiratory syndrome coronavirus 2 (SARS-CoV-2) infection, had been confirmed globally, with more than 2 million related deaths (https://covid19.who.int/). Although most individuals remain asymptomatic or display mild symptoms, 15-20% of patients exhibit severe symptoms, specifically respiratory distress, often requiring mechanical ventilation or/and intensive care (ICU) admission [1], with a mortality rate after ICU admission estimated around 40% [2]. Multiple studies have identified profound underlying conditions that demonstrate increased susceptibility to a more severe prognosis and a higher risk of fatality, including the male gender, old age [3], or certain underlying medical conditions, such as hypertension, cardiovascular diseases, diabetes or obesity [4]. Additionally, patients infected with SARS-CoV-2 present metabolic dysregulation, possibly due to immune-triggered inflammation or other changes in the host physiology, and that these alterations often reflect the disease severity [1,5,6]. For instance, levels of particular amino acids positively correlated with severe COVID-19 cases [1,7]. Moreover, perturbations in energy metabolisms, TCA and urea cycle [6] and lipid metabolism [1,8] are correlated to disease prognosis. Thus, it is essential to assemble a complete metabolic signature correlated to disease severity to identify a set of biomarkers strongly associated with the patient outcome, with the final goal of employing them for diagnostics and therapeutic purposes.

Our study retrospectively analyzes the metabolome profile of 75 COVID-19 patients with moderate and severe symptoms admitted to Policlinico di Milano (Lombardy region, Italy) following SARS-CoV-2 infection between March and April 2020. Italy was the first Western country to experience COVID-19 disease, and the Lombardy region was the epicenter of the Italian COVID-19 pandemic. This cohort shows a higher mortality rate compared to others; therefore, it represents a unique opportunity to investigate the underlying metabolic profiles of the first COVID-19 patients in Italy and to identify potential predictive biomarkers.

## Methods

Plasma samples from 75 COVID19 patients were collected at the moment of admission to Policlinico di Milano (Italy) from March to April 2020 and SARS-CoV-2 positivity was confirmed by PCR (Ethical Clearance No. 462-2020-bis). Patients were initially classified based on the type of ventilation received: Nasal cannula (n=25) and VentMask (n=25) (moderate cases) or CPAP (n=25) (severe cases).

Plasma untargeted metabolomics was performed by Global Metabolomics (HD4) in Metabolon, (USA) as previously described [9]. The metabolomics method is ISO 9001:2015 certified and the lab is accredited by the College of American Pathologist (CAP), USA. Clinical data was collected from the registries.

Logistic regression was used to model associations of each biomarker with COVID-19-related in-hospital mortality adjusting for age, gender, and body mass index (BMI). As BMI was missing for many patients, multivariate imputation by chained equations (MICE) was used (via the MICE R-package) to impute BMI. Imputation was done in two stages, first including no biomarkers, and then including those biomarkers that were found to be significant under the original imputation. The resulting pooled estimates and inference were obtained using Rubin’s rules. P-values were not adjusted in this analysis.

Metabolomics data were log2 transformed and plotted using histograms with normal distribution superimposed. R package LIMMA was applied for differential abundance analysis between different mask types (Nasal cannula/VentMask/CPAP), outcome (survivors/non-survivors), and severity (moderate/severe). Adjustment for multiple testing was assessed using false discovery rate (FDR) <0.05. Heatmap was built using the R package ComplexHeatmap. Uniform Manifold Approximation and Projection (UMAP) representations were done using the R package UMAP. Metabolites with variance equal to zero were removed and positive significant pairwise correlations after Bonferonni correction (Spearman, adjusted *p*<0.00001) were used for association analysis. The strength of the connections was evaluated by plotting distribution of correlation coefficients in a graphical network using igraph (https://igraph.org/python/). The network was compared to a random network with similar dimensions to validate the structure of the network to be not due to chance. Community detection was performed using Leiden algorithm (https://leidenalg.readthedocs.io/en/stable/index.html). For each community large enough (n>30), metabolite set enrichment analysis (MSEA) with KEGG and Metabolon terms via the Python module gseapy was performed. The average degree and clustering coefficient were calculated for each community. The final network was build using Cytoscape and biomarkers that were significantly associated with death were highlighted.

## Results and discussion

Among the 75 patients enrolled in this study, 24% (18/75) succumbed to death in the hospital (Supplementary Table 1) and a significantly higher rate of fatal outcomes was observed for severe cases compared to moderate cases (*p=*0.0218, Chi-square test). Among patients who died, comorbidities were nearly universally observed (94%, 17/18). Underlying conditions, like hypertension, were recorded for 46% (26/57) of the survivors and 50% (9/18) of the non-survivors, while cardiovascular diseases were significantly more common in non-survivors (*p*=0.0320, Fisher’s exact test) compared to survivors. Among the serological markers, lactate was significantly higher (*p*<0.0001, Mann-Whitney test) in non-survivors compared to survivors, confirming its tight link to disease severity [10].

**Table 1:** Characteristics of the study population.

|  | Survivors | Non-survivors | *p value* |
| --- | --- | --- | --- |
| *Number* | 57 | 18 |  |
| *Gender* |  |  | 0.1169 |
| - *Male (%)* | 17 (30.4%) | 9 (50%) |  |
| - *Female (%)* | 40 (69.4%) | 9 (50%) |  |
| *Age in years, median (IQR)* | 62 (52-73.5) | 76.5 (68.5-82) | 0.0013 |
| *BMI, median (IQR)\** | 26.3 (24.4-29.5) | 29.6 (27-32.2) | 0.1697 |
| *SpO2, median (IQR)\** | 97 (94.25-98) | 95 (89.25-98) | 0.0768 |
| *pH, median (IQR)\** | 7.48 (7.45-7.5) | 7.49 (7.45-7.53) | 0.6672 |
| *Lactate, median (IQR)\** | 0.9 (0.7-1.1) | 1.5 (1.1-1.8) | <0.0001 |
| *Comorbidities (%):* | 38 (66.7%) | 17 (94%) | 0.0297 |
| - *Hypertension* | 26 (46.4%) | 9 (50%) | 0.7913 |
| - *Diabetes* | 9 (16%) | 3 (16.7%) | NS |
| - *Lung disease* | 4 (7.1%) | 1 (5.6%) | NS |
| - *COPD* | 4 (7.1%) | 2 (11.1%) | 0.6257 |
| - *Obesity* | 8 (14.3%) | 6 (33%) | 0.0867 |
| - *Renal disease* | 4 (7.1%) | 3 (16.7%) | 0.3481 |
| - *Liver disease* | 1 (1.8%) | 1 (5.6%) | 0.4249 |
| - *Cardiovascular disease* | 7 (12.5%) | 7 (38.9%) | 0.0320 |
| - *Cerebrovascular disease* | 2 (3.6%) | 0 | NS |
| - *Others\*\** | 14 (24.6%) | 11 (61.1%) |  |
| *Number of comorbidities (listed above):* |  |  | 0.0047 |
| - *None* | 19 (33.4%) | 1 (5.6%) |  |
| - *One* | 16 (28%) | 3 (16.7%) |  |
| - *Two* | 9 (15.8%) | 6 (33%) |  |
| - *Three or more* | 13 (22.8%) | 8 (44.4%) |  |
| *Disease severity§:* |  |  | 0.0218 |
| - *Moderate* | 42 (73.7%) | 8 (44.4%) |  |
| - *Severe* | 15 (26.3%) | 10 (55.6%) |  |

\* Calculated from the available data
\*\* Gastrointestinal reflux, dyslipidemia, neoplasia, dementia
§ Based on the masks

In a logistic regression analysis (adjusted to age, gender, and BMI), 35 metabolites, among the >1000 tested, were significantly associated with COVID-19 mortality (*p*<0.05, unadjusted), among which 10 biomarkers were significant at the unadjusted 0.025 level (Figure 1A). Interestingly, cyclic adenosine monophosphate (cAMP) is significantly increased in non-survivors compared to survivors (OR: 7.4 95% CI 1.5 - 37). cAMP is a well-known intracellular messenger that functions as a regulator of various cellular activities, including cell growth and differentiation, gene transcription, protein expression, and is intimately involved in mitochondrial dynamics [11]. As cAMP plays a role in SARS-CoV-2 endocytosis in the initial phases of the infection [12], its involvement in disease progression is worthy of further investigations as a potential biomarker.

**Figure 1:**
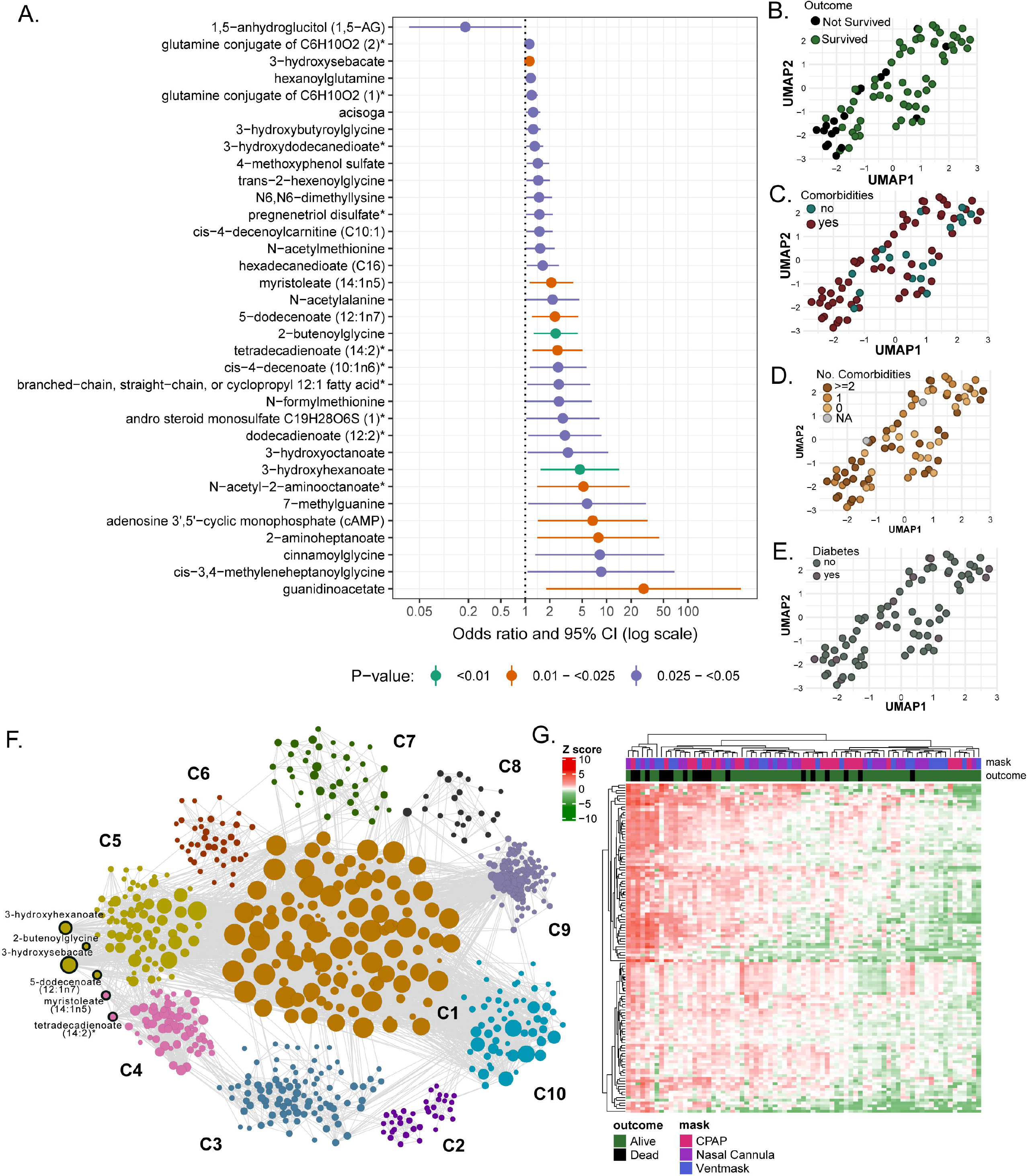
10 metabolites are significantly associated with COVID-19 related in-hospital mortality. A. All biomarkers that were significant at the 0.05 level, after adjustment of age, gender, and BMI, were included in the plot and order by the value of the odds ratio. Color coding for lower p-values is presented in the legend. B-E. UMAP visualization of patients’ data after selecting the 10 death biomarkers. Patients were labeled for the B. outcome, C. presence of comorbidities, D. amount of comorbidities and E. diabetes. F. Global weighted network after communities’ detection. Biomarkers with COVID-19 related mortality belong to communities 4 and 5 (labeled). The size of the bubble represents the connectivity of each metabolite. G. Heatmap of potential biomarkers and first neighbors in patients. Outcome (survivors/non-survivors) and mask type is indicated. Data were log-transformed.

Using UMAP, we observed that the distribution of the patients in the biomarker enrichment showed a separation between survivors and non-survivors (Figure 1B). No specific pattern or clustering was observed for comorbidities (Fig 1C), the number of comorbidities (Fig 1D), or diabetes (Fig 1E), indicating that these metabolite sets only differentiate related to the clinical outcome.

To further understand the patterns of metabolic changes related to COVID-19, we performed a weighted correlation network analysis on the metabolomics dataset, using significant positive correlations (Spearman, adjusted *p*<0.00001). We identified 10 metabolites communities highly connected in a network of 916 nodes (metabolites) and 11453 edges (Figure 1F). Six predicted biomarkers (out of the 10 previously identified with *p*<0.025, unadjusted) were highly correlated and were present in the network and belong to the lipid pathways. These metabolites are also known to be associated with peroxisomal fatty acid oxidation disorders (3-hydroxysebacate) [13] or insuline resistance (5-dodecenate (12:1n7), tetradecadienoate (14:2)* and myristoleate (14:1n5)) [14]. Finally, to further discriminate the subset of metabolites significantly associated with a fatal clinical outcome, we selected all the biomarkers and their first neighbors in the network analysis previously described (238 metabolites). Based on these data, we found a clear clustering of non-survivors in opposition to survivors (Fig 1G). No clustering according to mask type was observed, indicating that the metabolic signature associated with mortality appears to be independent of the oxygen demand at the moment of hospitalization, providing the first identified correlation between a metabolite profile and disease severity in COVID-19 patients.

It is important to consider that when this study was planned (March-April 2020), there was little knowledge about the COVID-19. Therefore, several clinical data were missing. Despite that, this is the first set of biomarkers identified from high throughput metabolomics data that are associated with mortality and are not confounded by other preexisting conditions.

## Conclusions

Our analysis has identified metabolic biomarkers that in our data differentiate between COVID-19 survivors and non-survivors and that may be predictive of death from COVID-19, from the early stage of the epidemic, independently from oxygen demand at the moment of diagnosis. Our results on high throughput metabolomics contribute to a better understanding of COVID-19-related metabolic disruption and may represent a useful starting point for the identification of independent prognostic factors to be employed in the therapeutic practice.

## Data Availability

All datasets generated for this study are available on request to the corresponding author.

## Authors’ contribution

U.N. and N.S.F. designed the study. E.S., A.B., M.Sciumè, F.M., A.A.L., S.A., M.Sachs, F.Billi, F.Blasi, E.G., G.C., P.D., S.K., A.G., F.P., L.S. analyzed the data and performed the statistical analyses. E.S., U.N., N.S.F. wrote the paper. U.N. and K.S. acquired the fundings. E.S, C.L., L.V., K.S., L.B., N.S.F., U.N. reviewed and edited the paper. All authors read and approved the final manuscript.

## Acknowledgments

The authors would also like to thank all patients involved in this study, as well as the dedicated medical and research staff who fight against SARS-CoV-2. We also like to acknowledge the work of the “COVID-19 NETWORK” working group:

Responsible person: Prof. Silvano Bosari Fondazione IRCCS Ca’ Granda Ospedale Maggiore Policlinico, Milan, Italy Scientific Direction: Silvano Bosari, Luigia Scudeller, Giuliana Fusetti, Laura Rusconi, Silvia Dell’Orto. Transfusion Medicine (Biobank): Daniele Prati, Luca Valenti, Silvia Giovannelli. Infectious Diseases Unit: Andrea Gori, Alessandra Bandera, Antonio Muscatello, Davide Mangioni, Laura Alagna, Giorgio Bozzi, Andrea Lombardi, Riccardo Ungaro, Teresa Itri, Valentina Ferroni, Valeria Pastore, Roberta Massafra, Ilaria Rondolini, Angelo Bianchi Bonomi. Internal Medicine, Hemophilia and Thrombosis Center and Fondazione Luigi Villa: Flora Peyvandi, Roberta Gualtierotti, Barbara Ferrari, Raffaella Rossio, Elisabetta Corona, Nicolò Rampi, Costanza Massimo. Internal Medicine, Immunology and Allergology: Nicola Montano, Barbara Vigone, Chiara Bellocchi, Giulia Coti, Mimma Sternativo. Respiratory Unit and Cystic Fibrosis Adult Center: Francesco Blasi, Stefano Aliberti, Maura Spotti, Edoardo Simonetta, Leonardo Terranova, Francesco Amati, Carmen Miele, Annalisa Vigni. Emergency Medicine: Giorgio Costantino, Monica Solbiati, Ludovico Furlan, Marta Mancarella, Giulia Colombo, Giorgio Colombo. Acute Internal Medicine: Valter Monzani, Angelo Rovellini, Filippo Billi, Christian Folli Internal Medicine: Marina Baldini, Irena Motta. General Medicine and Metabolic Diseases: Anna Fracanzani, Rosa Lombardi. Geriatric Unit: Matteo Cesari, Marco Proietti. Istituto di Ricerche Farmacologiche Mario Negri IRCCS: Alessandro Nobili, Mauro Tettamanti.

